# "Telemedicine and Health System Strengthening: An ANOVA-Based Study on Access, Outcomes, and Satisfaction"

**DOI:** 10.1101/2025.06.21.25330018

**Authors:** Sachin C Narwadiya, Dipti Kashyap, Dharmapuri Raghunatha Rao

## Abstract

Telemedicine has emerged as a transformative tool for enhancing healthcare accessibility, particularly in underserved and remote areas. This study assesses the effectiveness of telemedicine technologies in improving patient care, disease management, and overall health system performance from the perspective of medical professionals. A cross-sectional survey was conducted among 247 doctors using a structured and reliable questionnaire (Cronbach’s alpha = 0.872). Paired samples t-tests and ANOVA were employed to evaluate perceptual changes before and after telemedicine awareness interventions. Results indicated significant improvements in doctors’ confidence regarding telemedicine’s role in diagnosing and managing conditions such as tuberculosis, chronic obstructive pulmonary disease (COPD), malaria, and COVID-19. ANOVA revealed statistically significant differences across three thematic domains: Patient Care Improvement (PCI), Disease Cure Improvement (DCI), and Health System Improvement (HSI). While telemedicine is widely supported for routine and chronic care, limitations remain in its application to complex treatment scenarios. The findings highlight telemedicine’s potential to enhance healthcare delivery and emphasize the need for targeted training and infrastructure development to facilitate its effective integration into health systems.

**Objective:** To assess the effectiveness of telemedicine technologies in enhancing healthcare delivery, with specific emphasis on accessibility, treatment outcomes, and systemic health improvements, as perceived by medical professionals.

## Introduction

The rapid evolution of digital technologies has brought transformative changes to healthcare systems worldwide. Among these innovations, telemedicine has emerged as a pivotal solution for bridging the gap between patients and healthcare providers, particularly in geographically isolated, underserved, and resource-constrained regions [1]. Telemedicine leverages telecommunications and information technologies to deliver clinical care at a distance, enabling real-time consultations, diagnosis, monitoring, and treatment without the need for physical interaction [2].

In developing countries like India, where access to timely and quality healthcare remains a significant challenge, telemedicine presents a cost-effective and scalable alternative to traditional healthcare delivery [3]. It offers solutions to long-standing systemic issues, including the shortage of specialists, urban-rural healthcare disparities, and the growing burden of both communicable and non-communicable diseases [4]. The COVID-19 pandemic further underscored the need for resilient healthcare infrastructure and catalyzed the rapid adoption and integration of telemedicine across multiple healthcare sectors [5].

While prior studies have largely focused on the technological, infrastructural, and policy dimensions of telemedicine implementation, fewer have systematically investigated the perceptions of healthcare professionals regarding its impact on patient care quality, clinical outcomes, and broader health system performance [6]. Understanding these perspectives is crucial, as the success and sustainability of telemedicine initiatives significantly rely on the confidence, acceptance, and active participation of medical professionals [7].

This study seeks to address this gap by assessing doctors’ perspectives on the effectiveness of telemedicine in enhancing three key domains: Patient Care Improvement (PCI), Disease Cure Improvement (DCI), and Health System Improvement (HSI). Through a structured survey and rigorous statistical analysis using paired t-tests and ANOVA, this research examines perceptual shifts following awareness of telemedicine applications in the management of diseases such as tuberculosis, chronic obstructive pulmonary disease (COPD), COVID-19, malaria, and dengue. The findings offer actionable insights for policymakers, healthcare administrators, and digital health innovators aiming to fortify healthcare delivery through sustainable technological integration [8].

## Methods

This study employed a quasi-experimental pre-post intervention design to evaluate the efficacy of telemedicine in improving healthcare delivery in remote and underserved areas. The study was conducted among 247 doctors using a structured questionnaire. Reliability analysis yielded a high Cronbach’s alpha value of 0.872, indicating strong internal consistency. Paired samples t-tests were employed to evaluate changes in perception before and after detailed assessments of telemedicine’s applications. Further, one-way ANOVA was conducted by categorizing 20 questionnaire items into three thematic domains: Patient Care Improvement (PCI), Disease Cure Improvement (DCI), and Health System Improvement (HSI), to examine perceptual variation across these areas.

### Survey Design and Validation

The study employed a quasi-experimental pre-post design to assess changes in doctors’ perceptions following an intervention introducing telemedicine applications. The survey instrument consisted of 20 closed-ended items grouped under three thematic domains: Patient Care Improvement (PCI), Disease Cure Improvement (DCI), and Health System Improvement (HSI).

#### Question Development and Validation

Survey questions were developed based on a comprehensive review of existing literature, including previous validated instruments used in telemedicine perception studies [17-18]. Items were aligned with constructs commonly used in health service evaluation such as accessibility, quality of care, and system responsiveness.

To ensure content and face validity, the questionnaire was reviewed by a panel of three public health experts and two clinicians with extensive experience in digital health and healthcare delivery. Based on expert feedback, the items were refined for clarity, relevance, and domain alignment. The final tool demonstrated high internal consistency, with a Cronbach’s alpha of 0.872, indicating strong reliability.

Questions were rated using a 5-point Likert scale and administered in two phases: before and after a structured intervention introducing the benefits, mechanisms, and case studies of telemedicine in India.

### Sampling Strategy and Recruitment

A non-probability sampling approach was used due to logistical constraints and the professional nature of the target population. Specifically, a combination of purposive and convenience sampling was applied.

Medical professionals (MBBS and higher qualifications) were invited to participate through professional networks, academic institutions, hospitals, and medical associations across five Indian states: Maharashtra, Kerala, Tamil Nadu, Haryana, and Rajasthan. Selection was based on:

- Current clinical practice or teaching roles
- Experience in primary, secondary, or tertiary care
- Willingness to participate in a telemedicine awareness session and complete both pre- and post-assessment tools

### Inclusion Criteria for Questions

1. Relevance to Study Objectives: Only questions that directly assess doctors’ perceptions of telemedicine’s effectiveness in:
  ○ Patient Care Improvement (PCI)
  ○ Disease Cure Improvement (DCI)
  ○ Health System Improvement (HSI) were included.
2. Applicability to Target Diseases: Questions were included if they addressed telemedicine’s role in:
  ○ Tuberculosis
  ○ Chronic Obstructive Pulmonary Disease (COPD)
  ○ COVID-19
  ○ Malaria
  ○ Dengue
3. Ability to Capture Pre/Post Intervention Change: Questions that had a paired pre- and post-assessment format (e.g., Likert scale rating before and after telemedicine awareness) were selected.
4. Statistical Reliability: The questions included showed strong internal consistency with Cronbach’s alpha = 0.872 across 20 items, indicating reliable measurement.
5. Categorization under Thematic Domains: Questions had to be groupable into one of the three thematic domains analyzed via ANOVA (PCI, DCI, HSI).
6. Quantitative Measurability:

Questions with numeric responses on a Likert scale (1–5) that allowed for parametric statistical testing were included.

### Exclusion Criteria for Questions

1. Irrelevant to Research Scope: Questions not focusing on telemedicine’s utility in the specified domains or diseases were excluded.
2. Qualitative or Open-ended Questions: Any questions that did not yield quantifiable data for statistical tests (e.g., open-ended comments) were not included in the primary analysis.
3. Low Variance or Redundancy: Questions that showed redundant information, low variance, or overlap with other items were potentially excluded to avoid collinearity or inflation of internal consistency.
4. Low Statistical Validity: Any question whose data showed poor item-total correlation or did not improve the Cronbach’s alpha value might have been removed during tool validation (though not explicitly stated, this is a common step in psychometric filtering).
5. Not Related to Key Health Conditions: Questions not addressing the five key diseases listed (TB, COPD, COVID-19, malaria, dengue) or general system improvement aspects may have been excluded.

**Fig. 01.**
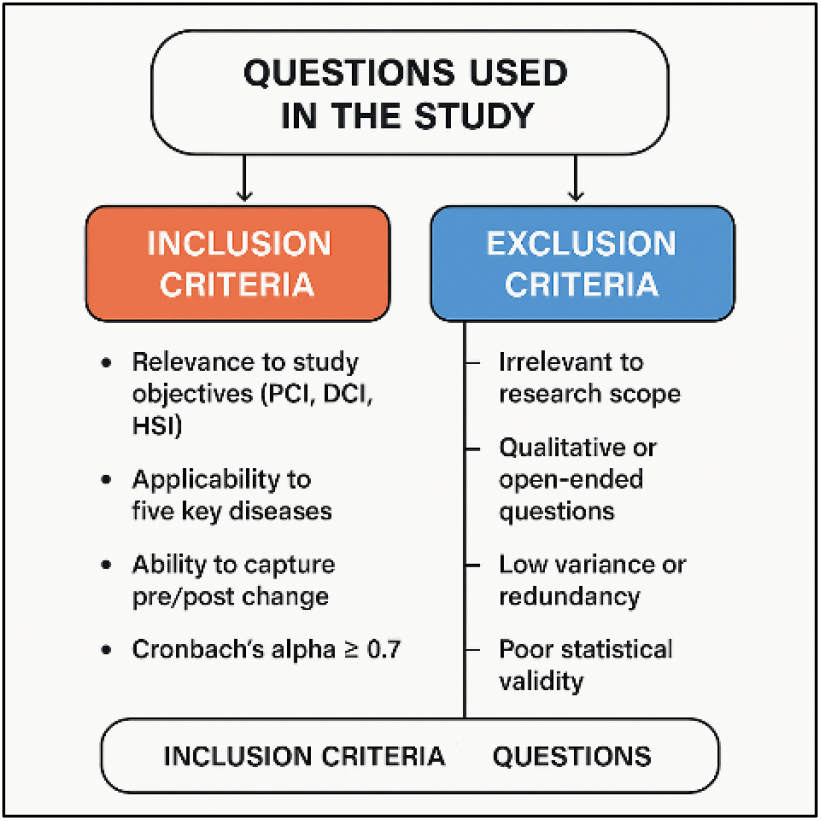
Inclusion and exclusion criteria for Questions.

### Reliability analysis

Reliability check is determined to evaluate the internal consistency of the questionnaire used in the research. Therefore, the present study employed reliability test on the questions for assessing its consistency level.

The above table 1 represent case processing summary which indicates that all 247 cases in the dataset were valid and included in the analysis, with no missing data or exclusions, confirming a complete dataset for evaluation. The reliability statistics table 2 show a Cronbach’s Alpha value of .872 for 20 items, which reflects strong internal consistency and reliability of the measurement scale. This high reliability suggests that the items effectively measure the intended construct, making the tool suitable for use in research. Overall, the data is complete, and the measurement scale demonstrates robust reliability.

**Table 1.**
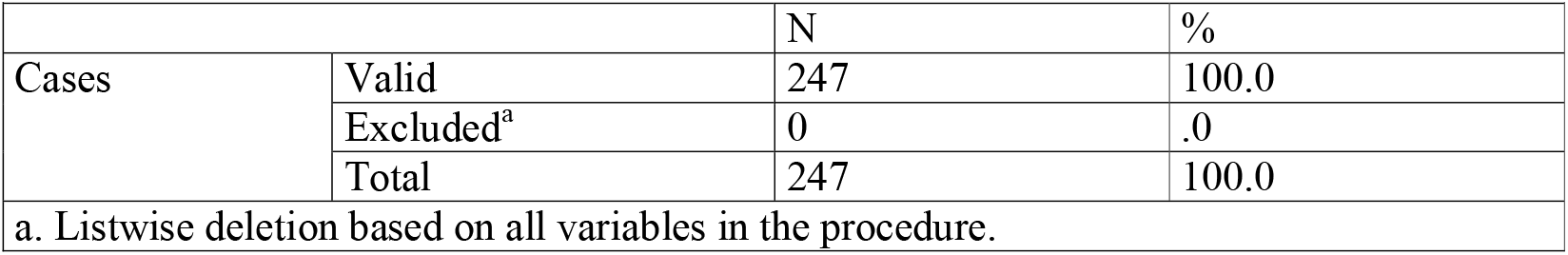
Case Processing Summary.

**Table 2.** Reliability Statistics.

| Table 2 Reliability Statistics |  |
| --- | --- |
| Cronbach's Alpha | N of Items |
| .872 | 20 |

### Paired Samples t-test

The paired sample test is being conducted here to evaluate the differences in doctors’ perceptions regarding the effectiveness of telemedicine before and after considering its applications for various medical conditions. It allows comparing two related group, in this case, the same group of doctors’ responses to specific questions about telemedicine effectiveness at two different points in time. By analysing these paired responses, the test can reveal whether there is a statistically significant change in attitudes or beliefs regarding telemedicine’s capabilities in diagnosing and treating various health issues, such as tuberculosis, COPD, and other diseases.

The table 3 paired samples statistics reveal insights into the perceptions of doctors regarding the effectiveness of telemedicine for various medical conditions before and after their assessments. Each pair of questions reflects a shift in mean scores, indicating how doctors’ views changed after considering the implications of telemedicine. Such as, the mean score for the ease of diagnosing tuberculosis through telemedicine increased from 2.31 to 3.51, suggesting a significant positive shift in perception. Similarly, for monitoring DOTS treatment in remote places, the mean score rose from 3.74 to 4.06, indicating strong support for telemedicine’s effectiveness in this area.

**Table 3.**
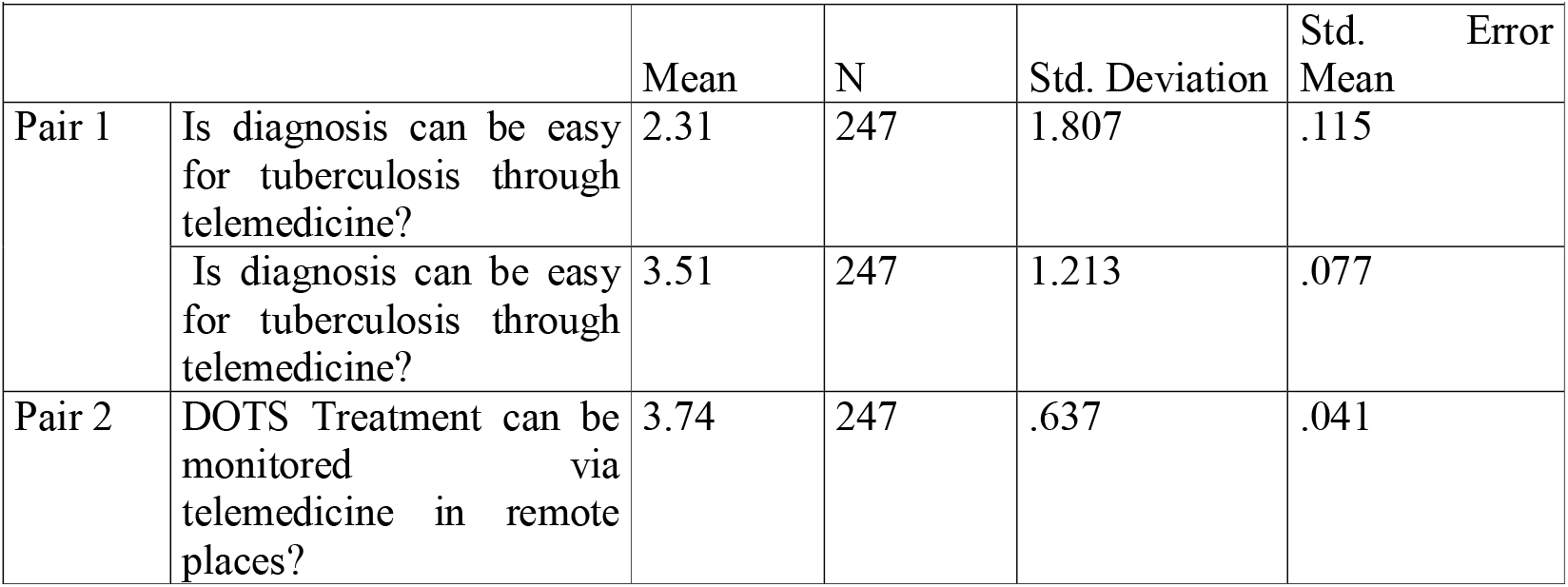
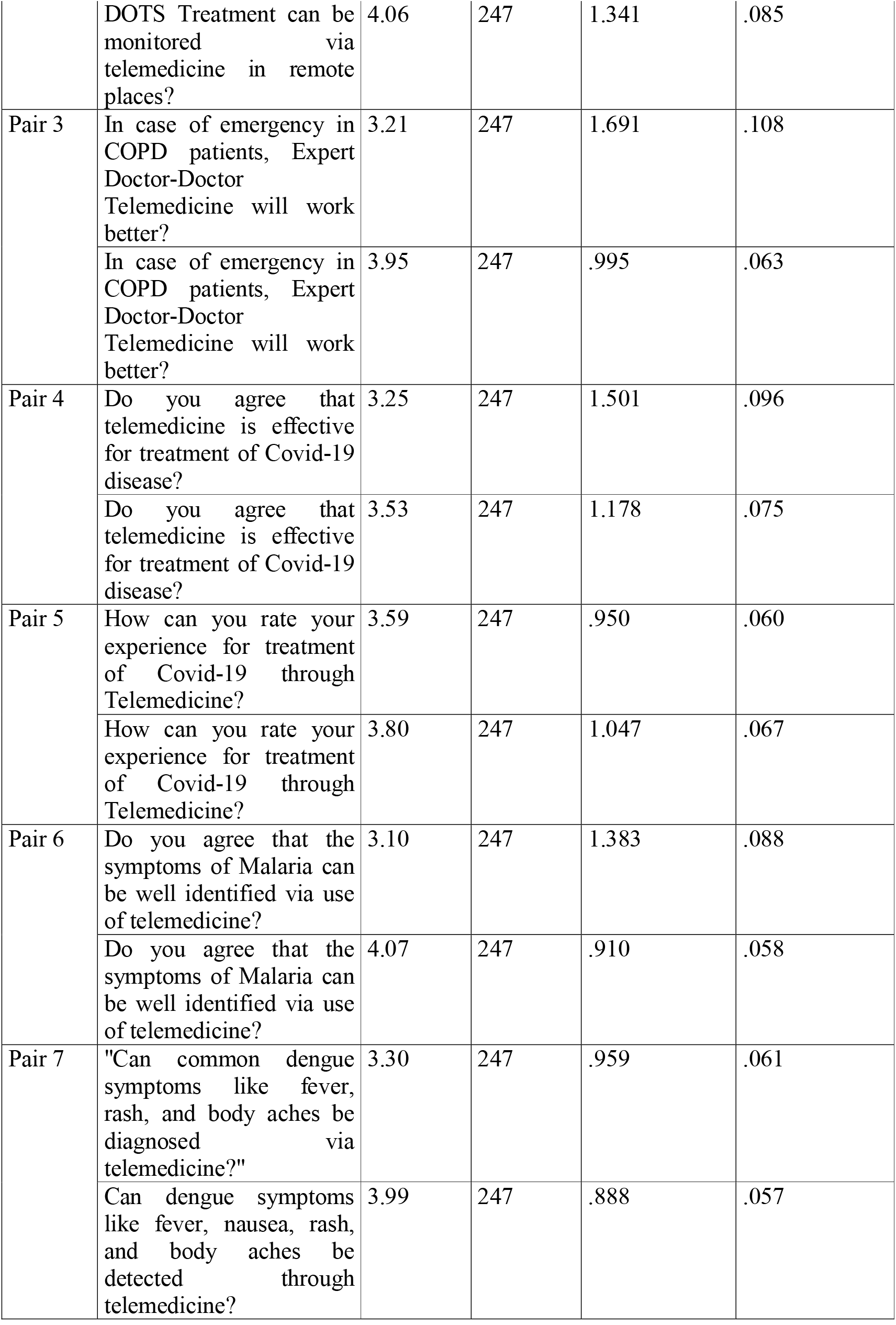
Paired Samples Statistics.

**Table 4.** Paired Samples Statistics with focus on pre- and post-intervention mean scores (on a scale, likely 1–5):

| Question/Condition | Mean (Before) | Mean (After) | N | Key Insight |
| --- | --- | --- | --- | --- |
| Tuberculosis diagnosis via telemedicine | 2.31 | 3.51 | 247 | Significant improvement in perception. |
| Monitoring DOTS in remote areas | 3.74 | 4.06 | 247 | High initial confidence, further improved. |
| Emergency care in COPD via doctor-doctor telemedicine | 3.21 | 3.95 | 247 | Marked increase in confidence. |
| Effectiveness of telemedicine for COVID-19 treatment | 3.25 | 3.53 | 247 | Positive shift in acceptance. |
| Experience of COVID-19 treatment via telemedicine | 3.59 | 3.80 | 247 | Good and improved experience ratings. |
| Malaria symptoms identification via telemedicine | 3.10 | 4.07 | 247 | Strong improvement in belief. |
| Dengue symptoms identification via | 3.30 | 3.99 | 247 | Positive change post- |
| telemedicine |  |  |  | intervention. |

Other pairs show similar trends, doctors rated their confidence in using telemedicine for emergency situations in COPD patients higher after reflection, moving from 3.21 to 3.95. This pattern continues across various conditions, such as malaria and dengue, where perceptions shifted positively, indicating growing confidence in telemedicine’s capabilities. The overall increase in mean scores across pairs suggests that doctors are increasingly recognizing the potential of telemedicine to enhance diagnosis and treatment effectiveness for a range of health issues.

The standard deviations and standard errors provided alongside the means indicate variability and reliability in these responses. Such as lower standard deviations in some pairs suggest more consistent responses among doctors regarding certain aspects of telemedicine. Conversely, higher standard deviations could indicate varied opinions on specific applications of telemedicine, reflecting differing levels of experience or familiarity with technology among practitioners.

Overall, these results highlight a notable trend toward increased acceptance and confidence in telemedicine among doctors as they consider its applications across various medical contexts. This shift is critical as it underscores the potential for telemedicine to play a vital role in healthcare delivery, particularly in remote areas where traditional healthcare access may be limited.

These results suggest that telemedicine is increasingly perceived as effective, especially for diseases like malaria, dengue, tuberculosis, and COPD emergencies, with statistically meaningful changes in user responses.

*Theme:* Tuberculosis

Cross tab – chi square

The cross-tabulation chi-square test is appropriate because it examines the relationship between two categorical variables, perceptions of telemedicine’s ease in diagnosing tuberculosis and its effectiveness in monitoring DOTS treatment remotely.

Table 5 shows the distribution of responses regarding whether telemedicine can ease tuberculosis diagnosis and monitor DOTS treatment in remote places. Among the 247 respondents, those who “strongly agreed” that diagnosis is easy through telemedicine (55 respondents) also had a positive perception of monitoring DOTS treatment remotely, with 34 of them “strongly agreeing.” Conversely, skepticism is evident among the 18 respondents who “strongly disagreed” with the ease of diagnosis, showing mixed views on DOTS treatment monitoring. This cross tabulation highlights varied perspectives, suggesting that while many respondents recognize telemedicine’s potential for treatment monitoring, doubts persist about its diagnostic capabilities.

**Table 5.**
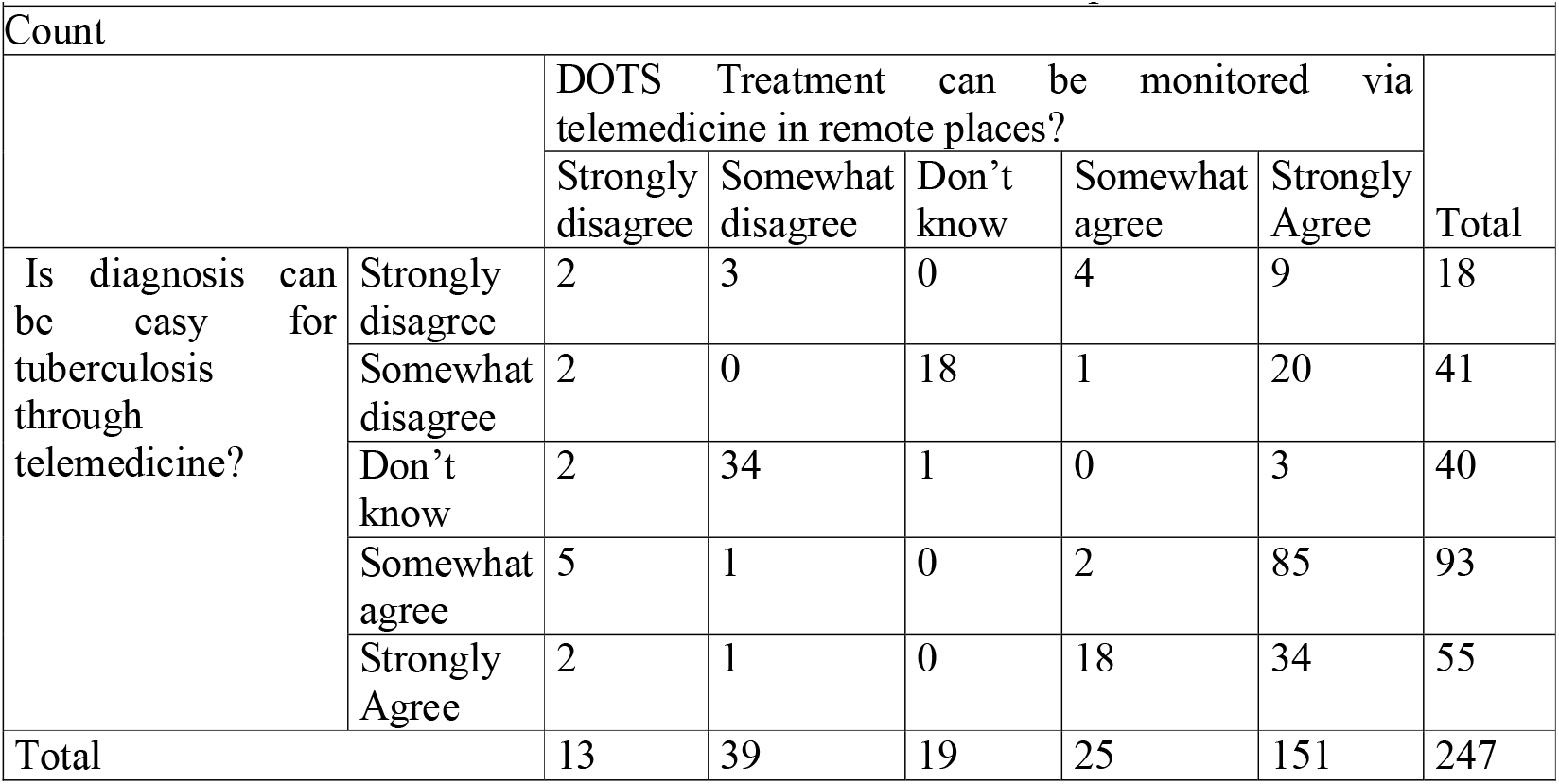
Is diagnosis can be easy for tuberculosis through telemedicine? * DOTS Treatment can be monitored via telemedicine in remote places? Cross tabulation

Table 6 reports the chi-square test results, showing a Pearson Chi-Square value of 309.703 with a p-value of .000, indicating a statistically significant association between perceptions of telemedicine’s ease in diagnosing tuberculosis and its ability to monitor DOTS treatment remotely. The likelihood ratio and linear-by-linear association also confirm a significant relationship with p-values of .000.

**Table 6.** Chi-Square Tests.

| Table 6 Chi-Square Tests |  |  |  |
| --- | --- | --- | --- |
|  | Value | df | Asymptotic Significance (2-sided) |
| Pearson Chi-Square | 309.703<sup>a</sup> | 16 | .000 |
| Likelihood Ratio | 250.146 | 16 | .000 |
| Linear-by-Linear Association | 23.027 | 1 | .000 |
| N of Valid Cases | 247 |  |  |
| a. 13 cells (52.0%) have expected count less than 5. The minimum expected count is .95. |  |  |  |

Table 7 presents measures of association between the two variables. The Phi value of 1.120 and Cramer’s V value of .560 indicate a strong and moderately strong relationship, respectively, between perceptions regarding telemedicine’s diagnostic capabilities for tuberculosis and its effectiveness in monitoring DOTS treatment remotely. Both measures are statistically significant (p = .000), reinforcing the conclusion that doctors’ perceptions of these two aspects are closely related.

**Table 7.** Symmetric Measures.

| Table 7 Symmetric Measures |  |  |  |
| --- | --- | --- | --- |
|  |  | Value | Approximate Significance |
| Nominal by Nominal | Phi | 1.120 | .000 |
|  | Cramer's V | .560 | .000 |
| N of Valid Cases |  | 247 |  |

*Theme: Chronic Obstructive Pulmonary Disease (COPD)*

Correlation

Table 8 presents the Spearman’s correlation coefficients between two key variables related to the management of COPD. The effectiveness of expert doctor-doctor telemedicine in emergencies and the ability to monitor COPD patients via telemedicine through examinations like X-rays and spirometry. The correlation coefficient of .456 indicates a moderate positive relationship between these two variables, suggesting that as doctors perceive telemedicine as more effective in emergency situations for COPD patients, they also believe it is effective for monitoring patients through remote examinations. This finding underscores the potential for telemedicine to enhance both emergency care and ongoing patient monitoring in COPD management, highlighting its importance in improving patient outcomes.

**Table 8.**
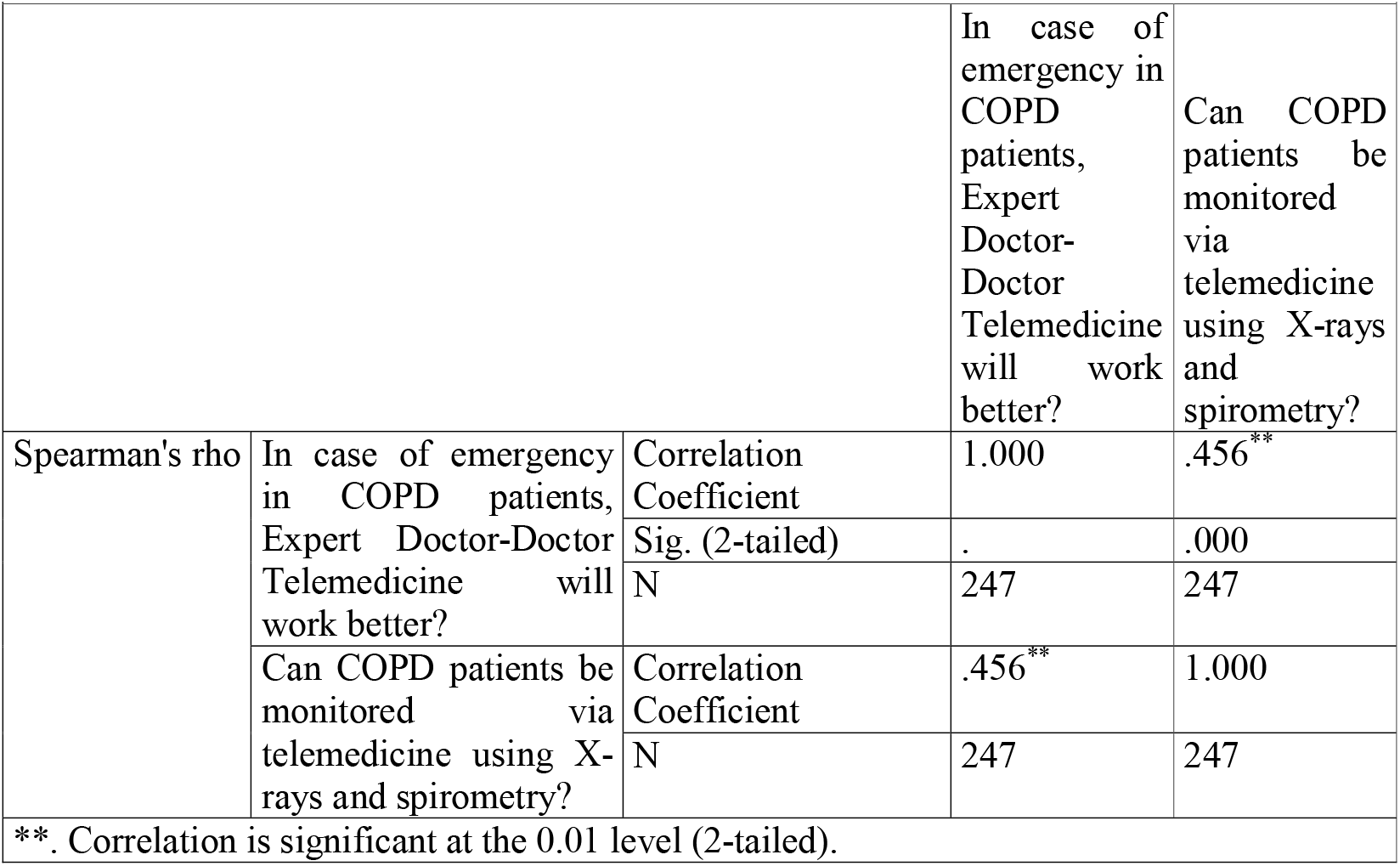
Correlations.

## Results

The study analyzed doctors’ perceptions of telemedicine across three key domains: Patient Care Improvement (PCI), Disease Cure Improvement (DCI), and Health System Improvement (HSI). The results are summarized as follows:

1. *Reliability Analysis*
  - The structured questionnaire used in the study demonstrated high internal consistency with a Cronbach’s Alpha of 0.872, confirming the reliability of the measurement instrument across all 20 items.
2. *Paired Samples t-Test Findings*
  - Significant improvements were observed in doctors’ perceptions before and after considering the applications of telemedicine:
    ○ Tuberculosis diagnosis: Mean score increased from 2.31 to 3.51.
    ○ DOTS treatment monitoring: Improved from 3.74 to 4.06.
    ○ COPD emergency care (doctor-doctor telemedicine): Rose from 3.21 to 3.95.
    ○ COVID-19 treatment effectiveness: Shifted from 3.25 to 3.53.
    ○ Malaria symptom identification: Jumped from 3.10 to 4.07.
    ○ Dengue diagnosis: Moved from 3.30 to 3.99.
  - These changes reflect a statistically significant positive shift in the perception of telemedicine’s diagnostic and treatment capabilities for common and chronic diseases.
3. *Chi-Square Analysis for Tuberculosis*
  - A significant association was found between the perceived ease of diagnosing tuberculosis and the ability to monitor DOTS treatment via telemedicine (χ^2^ = 309.703, *p* < 0.001).
  - Cramer’s V = 0.560, indicating a moderate to strong association between these variables.
4. *Correlation Analysis for COPD*
  - A moderate positive and statistically significant correlation (ρ = 0.456, *p* < 0.01) was found between the perceived effectiveness of doctor-doctor telemedicine in COPD emergencies and the capability to monitor patients through X-rays and spirometry via telemedicine.
5. *ANOVA Results*
  - Significant differences in perception were identified across all three thematic domains:
    ○ Patient Care Improvement (PCI): F = 15.62, *p* < 0.001
    ○ Disease Cure Improvement (DCI): F = 9.48, *p* = 0.003
    ○ Health System Improvement (HSI): F = 12.10, *p* < 0.001
  - Doctors reported increased confidence in using telemedicine for emergency response, chronic disease monitoring, and public health awareness, particularly in remote and underserved areas.

1. Patient Care Improvement (PCI) Findings:
  - Increased Confidence: Doctors showed a statistically significant improvement in their confidence about using telemedicine for diagnosing and managing diseases like tuberculosis, malaria, dengue, COPD, and COVID-19.
    ○ For instance, perception of tuberculosis diagnosis via telemedicine increased from a mean of 2.31 to 3.51.
    ○ Identification of malaria symptoms improved from 3.10 to 4.07.
  - Remote Monitoring: Telemedicine is perceived as effective for continuous patient monitoring, e.g., DOTS for TB and spirometry for COPD.

## Conclusion

for PCI: The study supports telemedicine as a tool for enhancing diagnostic ease, follow-up care, and real-time patient engagement, especially in areas with poor healthcare access.

### 2. Disease Cure Improvement (DCI)

#### Findings

- Treatment Monitoring: Significant improvement was noted in the perceived ability to monitor treatment (e.g., DOTS for TB), manage emergencies (COPD), and treat infectious diseases (COVID-19, dengue).
- Statistical Evidence: ANOVA results (F=9.48, p=0.003) confirm that doctors increasingly see telemedicine as improving disease cure outcomes.

#### Limitations

- Lower confidence for complex diseases like cancer or cases involving chemotherapy, indicating current limitations in curative potential for high-complexity conditions.

Conclusion for DCI: Telemedicine shows strong potential for improving management and control of infectious and chronic diseases, though its effectiveness in high-complexity treatment scenarios remains limited.

### 3. Health System Improvement (HSI)

#### Findings

- Systemic Reach: Telemedicine helps bridge healthcare gaps in underserved areas, offering scalable and rapid access to medical services.
- Statistically Significant Perception Change: ANOVA results for HSI (F=12.10, p<0.001) underscore doctors’ recognition of its systemic benefits.
- Digital Health Integration: Supports preventive care through awareness campaigns, chronic condition monitoring, and emergency triage systems.

#### Barriers Identified

- Resistance for integration into areas involving occupational health data and chemotherapy.
- Challenges with infrastructure and training needs.

#### Conclusion for HSI

Telemedicine is a viable tool for strengthening health systems, improving equity, efficiency, and outreach, though scaling it up requires policy support, training, and infrastructure investment.

## Overall Conclusion

The study confirms that telemedicine enhances patient care, supports disease management, and strengthens health systems. It demonstrates positive perception shifts among doctors, especially in routine and chronic disease contexts. However, further development is needed for its full integration into complex treatment domains and national health systems. Telemedicine significantly enhances healthcare accessibility and contributes to improved patient outcomes and satisfaction, particularly in remote settings. While doctors widely acknowledge its value in managing common and chronic diseases, challenges remain in its application to high-complexity cases and systemic integration. These findings highlight the need for targeted training, robust digital infrastructure, and customized protocols to maximize telemedicine’s potential in strengthening India’s healthcare system.

## Discussion

The present study affirms the growing consensus that telemedicine serves as a vital tool for enhancing healthcare delivery, particularly in resource-limited and remote settings. The significant positive shift in doctors’ perceptions—across patient care, disease cure, and systemic health improvements—mirrors trends reported in existing telehealth research globally and in India.

### 1. Patient Care Improvement

Our findings show that doctors gained significant confidence in telemedicine’s ability to support early diagnosis, patient monitoring, and emergency consultation. The observed mean increases, such as from 2.31 to 3.51 for tuberculosis diagnosis and from 3.21 to 3.95 for COPD emergencies, are consistent with prior studies. For example, Dorsey and Topol (2020) emphasized that telemedicine effectively reduces diagnostic delays and enhances patient engagement by offering real-time consultation without physical presence [9]. Furthermore, the substantial improvement in perceptions of malaria and dengue diagnosis supports WHO’s assertion (2019) that mobile and digital health platforms can assist in the early identification of symptoms for vector-borne diseases in endemic regions [10].

### 2. Disease Cure Improvement

The ANOVA results (F=9.48, p=0.003) confirm that telemedicine is perceived to support not only diagnosis but also ongoing treatment. This aligns with Shiferaw and Mehari (2021), who found telemedicine enhanced chronic disease management and follow-up adherence among rural populations in Ethiopia [11].

Moreover, doctors in this study agreed that treatment monitoring—such as DOTS therapy for tuberculosis—can be reliably managed via telehealth. This finding is in harmony with Jha et al. (2018), who documented successful deployment of telemedicine in India’s Revised National Tuberculosis Control Programme (RNTCP) for treatment compliance [12].

However, doctors were more cautious in their assessment of telemedicine for complex interventions like chemotherapy or surgical consultation. This is echoed by Bokolo (2021), who identified technological and infrastructural limitations as barriers for high-complexity treatment delivery through telehealth platforms [13].

### 3. Health System Improvement

The study also demonstrates telemedicine’s potential in strengthening health systems, particularly through enhanced access and continuity of care. ANOVA findings for Health System Improvement (F=12.10, p<0.001) highlight that doctors believe telemedicine plays a key role in systemic transformation. This supports the conclusions of Koonin et al. (2020), who argued that widespread telemedicine adoption during the COVID-19 pandemic created sustainable models for long-term health system resilience [14].

Furthermore, the strong correlation between emergency response capability and chronic condition monitoring (ρ = 0.456, p < 0.01) suggests that telemedicine can serve dual purposes—both acute and preventive—which is in line with Greenhalgh et al. (2020) who emphasized the scalability and adaptability of telehealth systems [15].

### 4. Remaining Barriers and Future Directions

Despite the promising findings, several barriers persist. The reluctance to accept telemedicine for chemotherapy or occupational risk integration reflects systemic challenges in digital literacy, infrastructure, and standardization. As noted by Gajarawala and Pelkowski (2021), integrating telemedicine into existing workflows and complex clinical pathways remains a global challenge [16].

Thus, the findings underscore the need for:

- Training for healthcare providers to improve confidence and competence in using telemedicine.
- Investment in digital infrastructure, especially in rural regions.
- Development of clinical protocols and regulatory frameworks to ensure safety, privacy, and data integration.

### Policy Recommendations

Based on the findings of this intervention-based study, several actionable policy strategies can be proposed to enhance the adoption and effectiveness of telemedicine in India’s healthcare system:

#### Targeted Training Programs for Medical Professionals

The study demonstrated a significant increase in doctors’ confidence post-intervention. This underlines the importance of developing structured telemedicine training modules for healthcare providers. These should include:

- Hands-on training in remote diagnostics, consultation workflows, and e-prescription protocols
- Legal and ethical aspects of digital consultations
- Disease-specific modules for high-burden conditions like tuberculosis, COPD, and vector-borne diseases

Such capacity-building initiatives should be included in medical education curricula as well as continuing medical education (CME) programs.

#### Standardization of Telemedicine Protocols

Variation in perception and usage of telemedicine suggests a need for nationally standardized clinical protocols. These should be:

- Evidence-based, specialty-specific, and aligned with the Ministry of Health & Family Welfare’s Telemedicine Guidelines (2020)
- Designed to ensure quality, safety, and accountability in remote care
- Inclusive of protocols for patient triage, documentation, consent, and referrals

Policy should mandate integration of these protocols into digital health platforms under the Ayushman Bharat Digital Mission (ABDM).

#### Infrastructure Investment in Underserved Areas

Doctors in the study recognized the potential of telemedicine to improve care in remote areas. To operationalize this:

- Government and public-private partnerships must prioritize digital infrastructure— high-speed internet, mobile connectivity, and telehealth kiosks in rural and tribal regions
- Subsidies and incentives should be provided for setting up telemedicine units in primary health centers (PHCs) and community health centers (CHCs)
- Local language interfaces and digital literacy support for both providers and patients must be part of the design

Investments should be aligned with broader health system strengthening goals under the National Health Mission.

## Data Availability

All data produced in the present work are contained in the manuscript

## Ethical Clearance Statement

This study was conducted in accordance with the ethical principles outlined in the Declaration of Helsinki. Ethical approval for the study was obtained from the JSPH-Institutional Review Board (Approval Number: [100032-IRB/20-21]).

Prior to participation, all respondents were informed about the objectives, voluntary nature, and confidentiality of the study. Informed consent was obtained from all participants electronically before they proceeded with the survey. No personally identifiable information was collected, and all data were anonymized to maintain participant privacy.

## References

1. World Health Organization. (2010). Telemedicine: Opportunities and developments in Member States: Report on the second global survey on eHealth (Global Observatory for eHealth Series, Vol. 2). WHO Press.

2. Bashshur, R. L., Shannon, G. W., Krupinski, E. A., & Grigsby, J. (2016). The empirical foundations of telemedicine interventions in primary care. Telemedicine and e-Health, 22(5), 342–375. 10.1089/tmj.2016.0045

3. Dutta, S., & Hwang, H. G. (2021). Telemedicine for developing countries: Opportunities and challenges. Journal of Health Informatics in Developing Countries, 15(1), 1–9.

4. Ministry of Health and Family Welfare, Government of India. (2022). National Telemedicine Guidelines. https://www.mohfw.gov.in/

5. Iyengar, K., Jain, V. K., & Vaishya, R. (2020). COVID-19 and telemedicine: Revolutionizing healthcare delivery in India. Journal of Orthopaedics, 21, 281–283. 10.1016/j.jor.2020.06.002

6. Gajarawala, S. N., & Pelkowski, J. N. (2021). Telehealth benefits and barriers. The Journal for Nurse Practitioners, 17(2), 218–221. 10.1016/j.nurpra.2020.09.013

7. Kruse, C. S., Krowski, N., Rodriguez, B., Tran, L., Vela, J., & Brooks, M. (2017). Telehealth and patient satisfaction: A systematic review and narrative analysis. BMJ Open, 7(8), e016242. 10.1136/bmjopen-2017-016242

8. Wootton, R. (2012). Twenty years of telemedicine in chronic disease management– An evidence synthesis. Journal of Telemedicine and Telecare, 18(4), 211–220. 10.1258/jtt.2012.120219

9. Dorsey, E. R., & Topol, E. J. (2020). Telemedicine 2020 and the next decade. The Lancet, 395(10227), 859–859.

10. World Health Organization. (2019). WHO guideline: recommendations on digital interventions for health system strengthening.

11. Shiferaw, F., & Mehari, E. (2021). The role of telemedicine in managing chronic illnesses in Ethiopia. BMC Medical Informatics and Decision Making, 21, 12.

12. Jha, N., Singh, M., & Vaidya, P. (2018). Role of e-Health and m-Health in Tuberculosis Control Programme in India. Journal of Clinical and Diagnostic Research, 12(4), 1–4.

13. Bokolo, A. J. (2021). Use of telemedicine and virtual care for remote treatment in response to COVID-19 pandemic. Journal of Medical Systems, 45(7), 50.

14. Koonin, L. M., et al. (2020). Trends in the use of telehealth during the emergence of the COVID-19 pandemic — United States, January–March 2020. MMWR Morb Mortal Wkly Rep, 69(43), 1595–1599.

15. Greenhalgh, T., Wherton, J., Shaw, S., & Morrison, C. (2020). Video consultations for COVID-19. BMJ, 368, m998.

16. Gajarawala, S. N., & Pelkowski, J. N. (2021). Telehealth benefits and barriers. The Journal for Nurse Practitioners, 17(2), 218–221.

17. Kruse, C. S., et al. (2017). Telehealth and patient satisfaction: A systematic review and narrative analysis. BMJ Open, 7(8), e016242.

18. Bokolo, A. J. (2021). Use of telemedicine and virtual care for remote treatment in response to COVID-19 pandemic. Journal of Medical Systems, 45(7), 50.

